# Associations of Skeletal Muscle Mass, Muscle Fat Infiltration, Mitochondrial Energetics, and Cardiorespiratory Fitness with Liver Fat Among Older Adults

**DOI:** 10.1101/2023.10.24.23297480

**Authors:** Daria Igudesman, Justine Mucinski, Stephanie Harrison, Peggy M. Cawthon, Jennifer Linge, Bret H. Goodpaster, Steven R. Cummings, Russell T. Hepple, Michael J. Jurczak, Stephen B. Kritchevsky, David Marcinek, Paul M. Coen, Karen D. Corbin

## Abstract

**Background:** Muscle mass loss may be associated with liver fat accumulation, yet scientific consensus is lacking and evidence in older adults is scant. It is unclear which muscle characteristics might contribute to this association in older adults.

**Methods:** We associated comprehensive muscle-related phenotypes including muscle mass normalized to body weight (D_3_-creatine dilution), muscle fat infiltration (MRI), carbohydrate-supported muscle mitochondrial maximal oxidative phosphorylation (respirometry), and cardiorespiratory fitness (VO_2_ peak) with liver fat among older adults. Linear regression models adjusted for age, gender, technician (respirometry only), daily minutes of moderate to vigorous physical activity, and prediabetes/diabetes status tested main effects and interactions of each independent variable with waist circumference (high: women—≥88 cm, men—≥102 cm) and gender.

**Results:** Among older adults aged 75 (IQR 73, 79 years; 59.8% women), muscle mass and liver fat were not associated overall but were positively associated among participants with a high waist circumference (β: 25.2; 95%CI 11.7, 40.4; *p*=.0002; N=362). Muscle fat infiltration and liver fat were positively associated (β: 15.2; 95%CI 6.8, 24.3; *p*=.0003; N=378). Carbohydrate-supported maximum oxidative phosphorylation and VO_2_ peak (adjusted β: −12.9; 95%CI −20.3, −4.8; *p*=0.003; N=361) were inversely associated with liver fat; adjustment attenuated the estimate for maximum oxidative phosphorylation although the point estimate remained negative (β: −4.0; 95%CI −11.6, 4.2; *p*=0.32; N=321).

**Conclusions:** Skeletal muscle-related characteristics are metabolically relevant factors linked to liver fat in older adults. Future research should confirm our results to determine whether trials targeting mechanisms common to liver and muscle fat accumulation are warranted.

## Introduction

Older adults are at an elevated risk of multiple metabolic derangements including liver fat accumulation. Excess liver fat is a component of non-alcoholic fatty liver disease (NAFLD) which is associated with an increased risk of cardiovascular events and mortality among persons of advanced age.^1^ Not only do older adults suffer from a higher prevalence of NAFLD (40% among the aging United States population) than their younger counterparts (25% in the general population^1,2^), but they are subject to a more aggressive disease prognosis once hepatic steatosis develops.^3^ Given the absence of ubiquitous clinical screening procedures for hepatic steatosis and the difficulty of accessing liver tissue for diagnosing NAFLD, it often goes undetected until more advanced stages of disease, such as advanced fibrosis, which can be irreversible and associated with poor outcomes.^4^ Thus, easily obtainable clinical measurements that are related to liver fat accumulation, are amenable to routine monitoring, and can serve as targets of lifestyle interventions to revert liver fat in older adults are urgently needed.

One such potential target which may provide insight into liver health is skeletal muscle mass. Although skeletal muscle mass and liver fat are inversely correlated in some studies— albeit most have not specifically focused on older adults—there is a lack of consensus in the extant literature about whether muscle loss is implicated in the pathophysiology of liver fat accumulation.^5,6^ In addition, no studies have comprehensively examined whether muscle characteristics such as myosteatosis, muscle mitochondrial energetics, and cardiorespiratory fitness (the whole body manifestation of muscle mass and mitochondrial energetics)^7,8^ may better reflect overall muscle health than muscle mass *per se*. Thus, it is tempting to speculate that ectopic lipid deposition and low oxidative capacity in muscle could drive muscle insulin resistance and downstream liver fat accumulation.^6,9–11^ Another potential reason for discordance of prior results is the variability in the rigor of measurements of muscle-related phenotypes and liver fat. Furthermore, despite the fact that numerous muscle characteristics are frequently compromised among older adults,^7,12,13^ it is unclear whether prior evidence in younger populations that link muscle phenotypes to liver fat can be generalized to older adults.

Given the lack of consensus regarding the connection between muscle mass and liver fat, the scarcity of such evidence focused on the older adult population, and a dearth of studies that have utilized rigorous measurement methods for both muscle and liver, our primary objective was to evaluate whether skeletal muscle mass was associated with liver fat using gold-standard measurements of both tissues in a sample of older adults in the Study of Muscle, Mobility and Aging (SOMMA) aged 75 (IQR 73, 79 years)—substantially older than earlier investigations (mean age ∼45-60 years).^14,15^ Secondarily, to explore beyond the link between muscle mass and liver fat and generate hypotheses about common mechanisms of pathobiology connecting the two tissues in the aging population, we interrogated the associations of muscle fat infiltration, muscle mitochondrial energetics, and VO_2_ peak with liver fat among SOMMA older adults. Recognizing that there are sex differences in both skeletal muscle mass^16^ and liver fat^17^ and that adiposity is a key risk factor for liver fat accumulation^1^ which may compromise the quality of skeletal muscle mass,^5,14^ we tested for effect modification by gender and waist circumference (a proxy for visceral adiposity)^18^ for all associations.

## Methods

### Study Participants

Older adults aged ≥70 years were recruited for SOMMA (https://www.sommastudy.com/) at the University of Pittsburgh and Wake Forest University School of Medicine between April 2019 and December 2021, as previously described.^19^ Briefly, individuals were eligible to participate if they were willing and able to complete a skeletal muscle biopsy and undergo MRI and magnetic resonance spectroscopy (MRS). Individuals were excluded if they reported an inability to walk one-quarter of a mile or climb a flight of stairs, had body mass index (BMI) ≥40 kg/m^2^, had an active malignancy, dementia, or any medical contraindication to biopsy or MR. Variables included in the present analysis were measured over three days of assessments at the baseline SOMMA visit: Day 1 (400 meter walk plus most other in-person assessments; Day 2 (cardiopulmonary exercise testing [CPET] and MR); and Day 3 (muscle biopsy). Participants self-reported their demographics (gender, race, and ethnicity), clinical data (health history including smoking, alcohol use, and medications), and lifestyle behaviors (self-reported diet and objective measures of physical activity). A binary variable (yes/no) was constructed for prediabetes or diabetes status based upon self-reported diabetes diagnosis or a hemoglobin A1c ≥5.7% measured at each study site’s local clinical laboratory (Local Quest Diagnostic Laboratory in Pittsburgh). Participants were asked to fast for 12 hours prior to the Day 3 baseline assessment for biospecimen collection (including blood, urine, and muscle).

### MRI

A whole-body MRI (Siemens Medical System 3T–Prisma [Pittsburgh] or Skyra [Wake Forest]) was used to assess body composition including quadriceps and total thigh muscle volume, muscle fat infiltration (mean fat fraction in the muscle tissue of the right and left anterior thighs), liver fat by proton density fat fraction (the ratio of the density of protons from triglycerides and the total density of protons from triglycerides and water expressed as a percentage^20^), and intra-abdominal (visceral) and abdominal subcutaneous adipose tissue (AMRA Medical, Sweden). For 3 participants with missing muscle fat infiltration data for the right leg, the left thigh fat infiltration was used instead of the mean of both legs. To compare the characteristics of participants with and without hepatic steatosis, continuous liver fat was dichotomized according to the threshold of ≥5.6 percent.^21^ However, liver fat was used as a continuous variable in all statistical models.

### Muscle Mass and Muscle Mitochondrial Energetics

The deuterium-labeled creatine (D_3_-creatine) dilution method was used to measure whole body muscle mass (D_3_Cr muscle mass) through assessment of creatine pool size, as previously described.^19^ D_3_Cr muscle mass was expressed as a percentage of total body weight in kg (i.e., D_3_Cr muscle mass/wgt).

A skeletal muscle biopsy was performed in the morning while the participant was fasted (12hrs) and had avoided strenuous physical activity for the prior 48 hours.^19^ Mitochondrial respiration was measured *ex vivo* in permeabilized muscle fibers using an Oxygraph O2K instrument (Oroboros Inc., Innsbruck, Austria), as previously described.^19^ In line with the hypothesis that greater carbohydrate-supported OXPHOS in peripheral tissues prevents liver fat accumulation,^22^ we focused on carbohydrate-supported maximal complex I and II-supported oxidative phosphorylation (Max OXPHOS, pmol/(s*mg). Fatty acid-supported OXPHOS with and without carbohydrate-supported OXPHOS was included in post-hoc correlation analyses with liver fat. Steady-state O_2_ flux was normalized to muscle fiber bundle wet weight using Datlab 7.4 software.

As an alternate measure of muscle mitochondrial energetics, we included maximal mitochondrial ATP production (ATP_max_, mM/second) measured *in vivo* by ^31^Phosphorus (^31^P) MRS of the quadriceps muscle following an acute bout of knee extensor exercise, as described elsewhere.^19^ A 3 Tesla MR magnet (Siemen’s Medical System—Prisma [Pittsburgh] or Skyra [Wake Forest]) collected the phosphorous spectra through the quadriceps. jMRUI software v6.0 was used to calculate ATP_max_.^19^

### Cardiorespiratory Fitness by CPET

A standardized treadmill CPET was used to measure cardiorespiratory fitness (VO_2_ peak). A modified Balke or manual protocol was used to measure oxygen consumption and carbon dioxide production during maximal effort exercise.^19^ VO_2_ peak was normalized to body weight (mL/kg/min).

### Anthropometrics

Body weight was measured in kg using a digital scale. Two to three measurements of waist circumference were taken using standardized procedures; the two closest measurements were averaged. Waist circumference was dichotomized as high (≥88 cm for women or ≥102 cm for men) or normal (<88 cm for women or <102 cm for men).^23^

### Moderate to Vigorous Physical Activity by Actigraphy

Participants wore the Actigraph GT9X (Actigraph, Pensacola, FL) on their non-dominant wrist for up to 7 days, as previously described.^19^ The data were scored using the ActiLife Software and used to calculate daily sedentary and active time in each 24-hr interval (midnight to 23:59).

### Statistical Analysis

We used descriptive statistics [mean ± SD or median (IQR) appropriate for normally and non-normally distributed continuous variables, respectively; and N (%) for categorical variables] to describe the demographic and clinical characteristics of the study sample. We restricted to participants with liver fat data and excluded the Wake Forest study site where data were unavailable due to logistical data collection issues (Participant Flow Diagram with additional details of participant inclusion and exclusion shown in **Supplementary Figure 1**). To evaluate the representativeness of our SOMMA subsample, we compared the characteristics of SOMMA participants included and excluded from our analysis.

We used general linear regression models to evaluate associations of an SD increment in each independent variable (primary: D_3_Cr muscle mass/wgt; exploratory: muscle fat infiltration, carbohydrate-supported *ex vivo* and *in vivo* mitochondrial bioenergetics, and cardiorespiratory fitness) with the outcome of liver fat. We evaluated linearity and model assumptions through inspection of the shape of each association and residual distributions, which led us to log-transform liver fat. To facilitate interpretability, we back-transformed model β estimates and 95% confidence intervals; this provided a proportional interpretation (i.e., the relative difference in liver fat associated with each SD increment in the independent variable). Model 1 was unadjusted; Model 2 was adjusted for age and gender (and technician when Max OXPHOS was the independent variable); and Model 3 was further adjusted for mean daily minutes of moderate to vigorous physical activity (MVPA) and prediabetes or diabetes status (**Supplementary Figure 2**). Given that self-reported use of anti-hyperglycemic agents (binary— yes/no) overlapped almost completely with prediabetes/diabetes status and only 58 participants in our sample reported using any such medication, we opted not to include medication use in statistical models.

We tested for effect modification between each independent variable with gender and waist circumference (a proxy for visceral adiposity^18^) for the outcome of liver fat by adding an interaction term between the independent variable and effect modifier to Model 3. We did not test for effect modification by visceral adipose tissue given that liver fat—the outcome variable— is a component of visceral adipose. We stratified results when the interaction term had a p-value <.1. All other results were deemed to be statistically significant at an α level of p<.05.

In post-hoc analyses, we examined whether fatty acid supported OXPHOS with or without carbohydrate-supported OXPHOS was correlated with log-transformed liver fat using the Spearman correlation coefficient. To gain further clinical insight with respect to NAFLD (liver fat percentage ≥5.6% or not), we included this as a binary outcome in exploratory analyses using logistic regression models with the same covariate structure and effect modifiers as the primary analysis. We conducted this supplementary analysis given the clinical relevance of NAFLD to the health of older adults. All analyses were performed using SAS version 9.4 (Cary, NC).

## Results

### Participant Characteristics

We included between 321 and 378 SOMMA participants in the analysis, based on data availability for each independent variable. Median age was 75 (IQR 73, 79 years), 59.8% were women, and 86.5% identified with a White race (**Table 1**). Median liver fat was 2.8% (IQR 2.0, 5.4%; range 1.0, 25.3%; N=378), 93 (24.6%) participants had hepatic steatosis, and mean D_3_Cr muscle mass/wgt was 28.7 ± 6.8% (N=362). SOMMA participants included in our analysis had a slightly higher BMI and lower hemoglobin A1c and VO_2_ peak than those excluded (**Supplementary Table 1**). A smaller proportion of included participants had prediabetes or diabetes (37.3%) as compared to those excluded (47.2%; *p*=.003).

**Table 1.** Baseline Characteristics of Participants With and Without Hepatic Steatosis.

|  |  |  |  |  |  |  |  |
| --- | --- | --- | --- | --- | --- | --- | --- |
|  | All Participants |  | NAFLD<sup>b</sup><br>(Liver Fat ≥5.6%) |  | No NAFLD<br>(Liver Fat <5.6%) |  |  |
|  | Total N | Mean ± SD,<br>Median (IQR), or N<br>(%)<sup>a</sup> | N | Mean ± SD,<br>Median (IQR),<br>or N (%) | N | Mean ± SD,<br>Median (IQR), or<br>N (%) | P-value |
| Demographics |  |  |  |  |  |  |  |
| Age (years) | 378 | 75 (73, 79) | 93 | 75 (73, 79) | 285 | 75 (73, 80) | 0.92 |
| Women | 378 | 226 (59.8) | 93 | 59 (63.4) | 285 | 167 (58.6) | 0.41 |
| Race | 378 |  | 93 |  | 285 |  | 0.81 |
| Asian |  | <5<sup>c</sup> |  | <5 |  | <5 |  |
| Black or African American |  | 45 (11.9) |  | 13 (14.0) |  | 32 (11.2) |  |
| Native American/Alaskan<br>Native |  | <5 |  | <5 |  | <5 |  |
| Other race/unknown |  | <5 |  | <5 |  | <5 |  |
| White or Caucasian |  | 327 (86.5) |  | 79 (85.0) |  | 248 (87.0) |  |
| Hispanic ethnicity | 378 | <5 | 93 | <5 | 285 | <5 | 0.57 |
| Clinical characteristics |  |  |  |  |  |  |  |
| Body mass index (kg/m<sup>2</sup>) | 378 | 27.8 ± 4.8 | 93 | 31 ± 4.4 | 285 | 26.7 ± 4.5 | **<0.0001** |
| Waist circumference<sup>d</sup> | 372 |  |  |  |  |  | **<0.0001** |
| Normal |  | 205 (55.1) |  | 23 (25.3) |  | 182 (64.8) |  |
| High |  | 167 (44.9) |  | 68 (74.7) |  | 99 (35.2) |  |

|  | All Participants |  | NAFLD<sup>b</sup><br>(Liver Fat ≥5.6%) |  | No NAFLD<br>(Liver Fat <5.6%) |  |  |
| --- | --- | --- | --- | --- | --- | --- | --- |
|  | Total N | Mean ± SD,<br>Median (IQR), or N<br>(%)<sup>a</sup> | N | Mean ± SD,<br>Median (IQR),<br>or N (%) | N | Mean ± SD,<br>Median (IQR), or<br>N (%) | P-value |
| Prediabetes or diabetes<sup>e</sup> | 378 | 141 (37.3) | 93 | 52 (55.9) | 285 | 89 (31.2) | **<0.0001** |
| Hemoglobin A1c (%) | 373 | 5.5 (5.3, 5.8) | 90 | 5.7 (5.5, 6.4) | 283 | 5.5 (5.2, 5.7) | **<0.0001** |
| Moderate to vigorous physical<br>activity (daily minutes) | 378 | 188 ± 87 | 93 | 179 ± 81 | 285 | 191 ± 89 | 0.24 |
| Outcome variable |  |  |  |  |  |  |  |
| Liver fat (%) | 378 | 2.8 (2.0, 5.4) | 93 | 8.1 (6.4, 12.1) | 285 | 2.4 (1.8, 3.1) | **<0.0001** |
| Exposure variables |  |  |  |  |  |  |  |
| Muscle mass (%) | 362 | 28.7 ± 6.8 | 89 | 28.2 ± 6.4 | 273 | 28.8 ± 7.0 | 0.48 |
| Muscle fat infiltration (%) | 378 | 7.3 (6.0, 9.1) | 93 | 8.4 (6.9, 10.0) | 285 | 7.1 (5.8, 8.7) | **<0.0001** |
| Carbohydrate-supported<br>maximum oxidative<br>phosphorylation<br>(pmol/(s*mg)) | 321 | 59.7 ± 19.2 | 78 | 57.9 ± 17.3 | 243 | 60.3 ± 19.8 | 0.34 |
| ATP production (mM/sec) | 345 | 0.52 ± 0.14 | 86 | 0.51 ± 0.13 | 259 | 0.52 ± 0.15 | 0.32 |
| VO<sub>2</sub> peak (mg/kg/min)<sup>f</sup> | 361 | 20.9 ± 5.4 | 85 | 19.1 ± 4.1 | 276 | 21.5 ± 5.6 | **<0.0001** |

<sup>a</sup> Note. Continuous variables presented as mean ± SD or median (IQR) when normally or non-normally distributed, respectively. Categorical variables presented as N (%). For continuous variables, p-values shown are from t-tests for normally distributed variables and from Wilcoxon rank sum tests for non-normally distributed variables. P-values for categorical variables are from the Chi-squared

|  | All Participants |  | NAFLD<sup>b</sup><br>(Liver Fat ≥5.6%) |  | No NAFLD<br>(Liver Fat <5.6%) |  |  |
| --- | --- | --- | --- | --- | --- | --- | --- |
|  | Total N | Mean ± SD,<br>Median (IQR), or N<br>(%)<sup>a</sup> | N | Mean ± SD,<br>Median (IQR),<br>or N (%) | N | Mean ± SD,<br>Median (IQR), or<br>N (%) | P-value |

### D_3_Cr Muscle Mass is Positively Associated with Liver Fat Only in Older Adults with a High Waist Circumference

Lower muscle mass is associated with a higher prevalence of hepatic steatosis in some, but not all studies.^5,6^ Therefore, we first asked whether higher muscle mass is associated with lower liver fat in an older study sample using the rigorous D_3_Cr muscle mass measurement and MRI proton density fat fraction for liver fat. Neither the crude nor the adjusted associations between D_3_Cr muscle mass/wgt and liver fat were statistically significant (fully adjusted β: −0.87, 95%CI: −8.1, 6.9, *p*=.82; **Table 2**, **Figure 1A**, **Supplementary Figure 1A**). However, the fully adjusted model indicated a positive association between D_3_Cr muscle mass/wgt and liver fat among participants with a high waist circumference (*p*=.001 for interaction; β: 25.2, 95%CI: 11.7, 40.4, *p*=.0002; N=160; **Figure 1B**, **Supplementary Figure 1B**), but not among those with a normal waist circumference (β: −3.6, 95%CI: −10.9, 4.4, *p*=.37; N=196).

**Figure 1.**
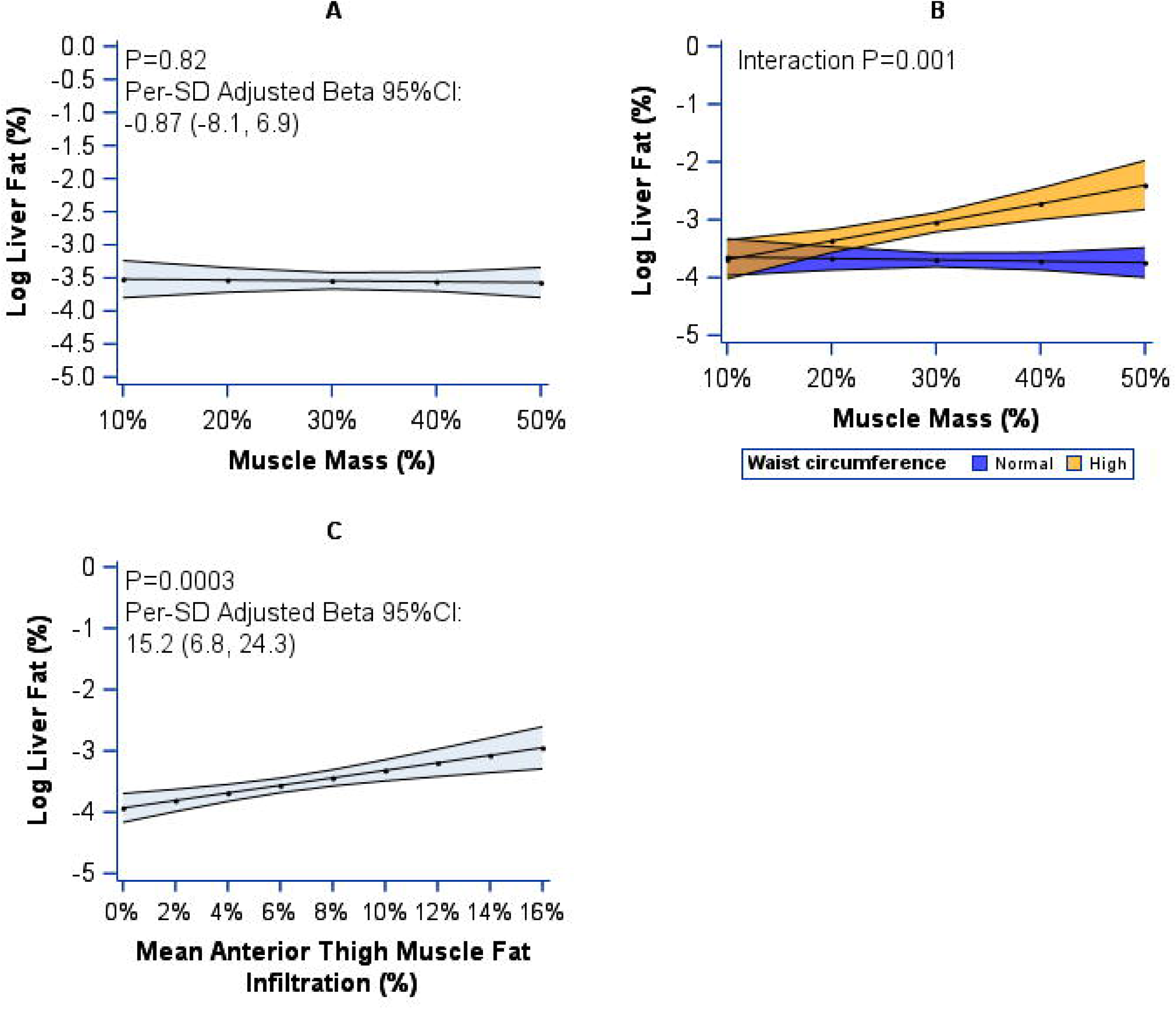
Associations of D_3_Cr Muscle Mass and Muscle Fat Infiltration With Liver Fat Among SOMMA Older Adults Panel (A) shows the association between each SD increment in muscle mass/wgt and log-transformed liver fat adjusted for age, gender, daily minutes of moderate to vigorous physical activity, and prediabetes/diabetes status (N=362). Panel (B) shows the association in panel (A) stratified by waist circumference (N=357). Panel (C) shows the fully adjusted association of muscle fat infiltration with liver fat (N=378). A high waist circumference was defined as ≥88 cm for women and ≥102 cm for men.

**Table 2.**
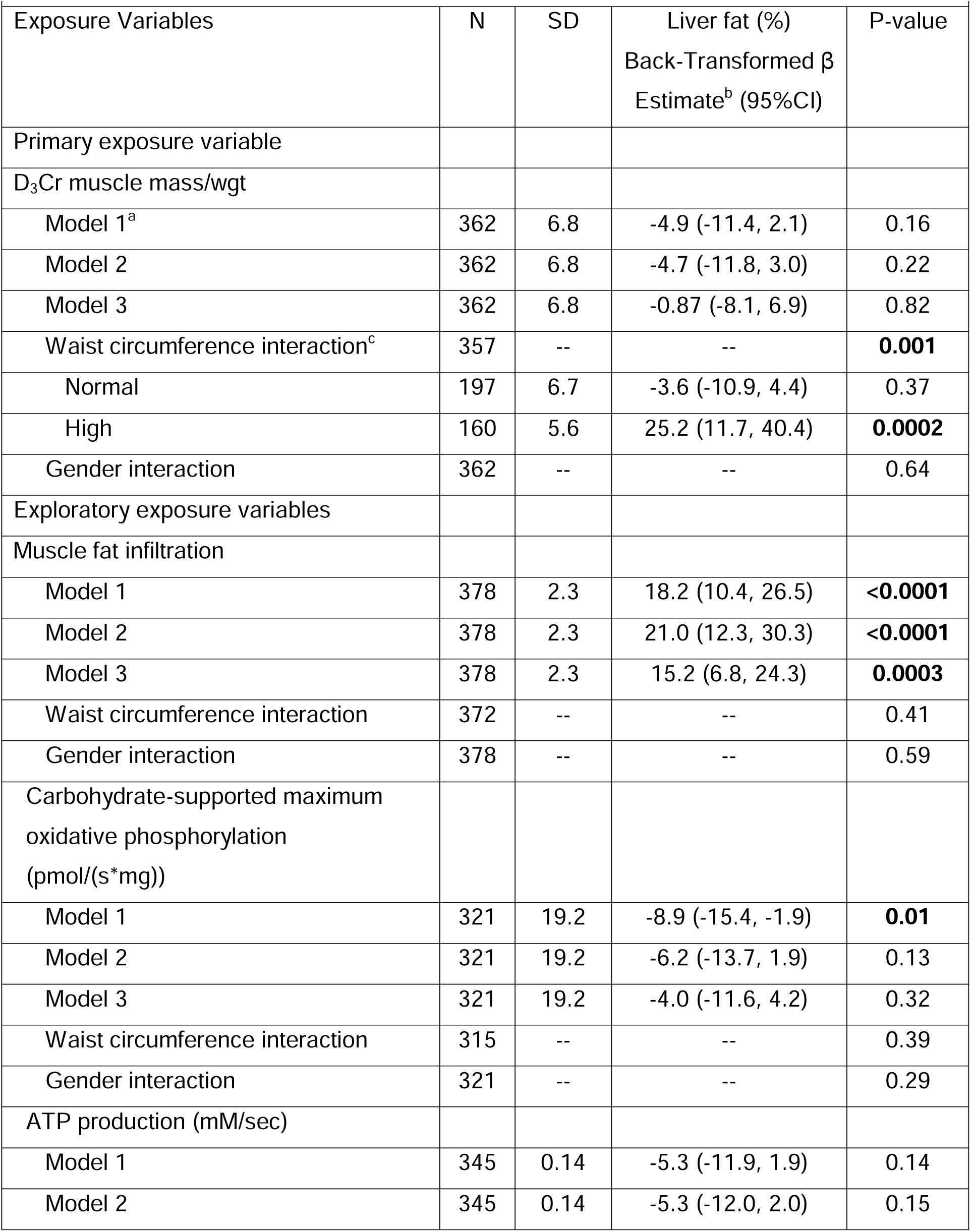
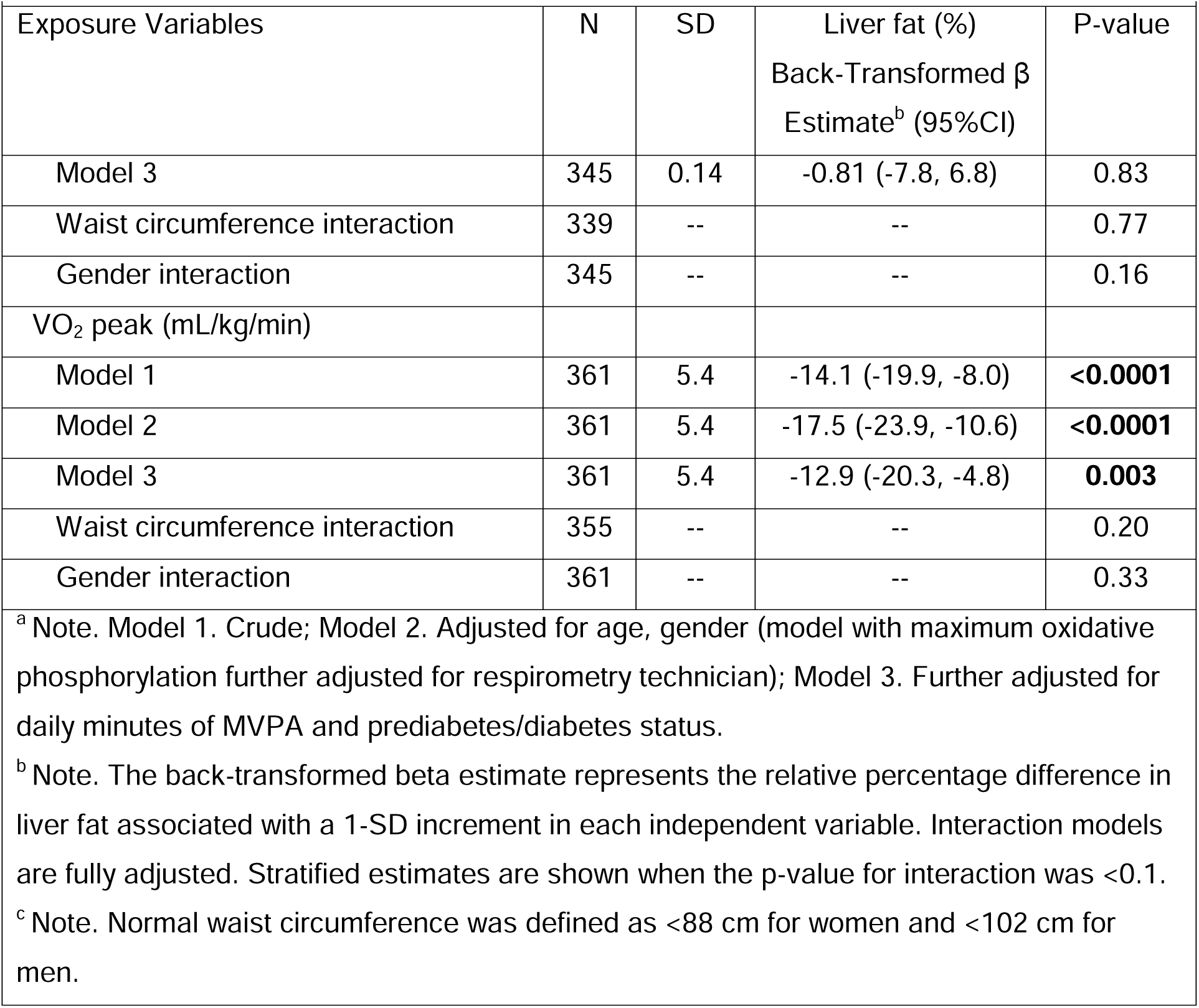
Associations of D_3_Cr Muscle Mass/Wgt, Muscle Characteristics, and Cardiorespiratory Fitness With Liver Fat.

### Skeletal Muscle Characteristics Including Muscle Fat Infiltration and Mitochondrial Bioenergetics are Associated with Liver Fat

Recent reports indicate that the skeletal muscle characteristic of estimated muscle fat infiltration is associated with liver fat in participants with advanced fatty liver disease and class III obesity.^10^ Through exploration of this association in an older sample, we uncovered a positive association according to the fully-adjusted model (15.2% higher liver fat per SD increment in muscle fat infiltration; 95%CI: 6.8, 24.3, *p*=.0003; N=378; **Figure 1C**, **Supplementary Figure 1C**) that did not vary by gender or waist circumference.

Lower mitochondrial energetics is a salient and modifiable skeletal muscle characteristic that is related to cardiometabolic disease,^8^ yet its association with liver fat is unclear. We showed for the first time an inverse association between *ex vivo* carbohydrate-supported muscle mitochondrial bioenergetics (Max OXPHOS) with liver fat according to the crude model (β: −8.9; 95%CI: −15.4, −1.9; *p*=0.01) but was attenuated to non-significance after adjustment for age, gender, and technician and was further attenuated in magnitude in the fully adjusted model (β: −4.0; 95%CI: −11.6, 4.2; *p*=.32; N=321; **Figure 2A**, **Supplementary Figure 1D**). Our post hoc exploration revealed a null crude correlation between fatty acid supported OXPHOS and liver fat (r=-0.006, *p*=.92; data not shown). This was also true for the combination of fatty acid and complex I or complex I&II carbohydrate-supported OXPHOS when correlated with liver fat (r=0.07, *p*=.26-.27; data not shown). ATP_max_, a secondary measure of muscle mitochondrial bioenergetics measured *in vivo*, was not associated with liver fat in either crude or adjusted models in the overall sample (fully adjusted β: −0.81; 95%CI −7.8, 6.8; *p*=.83; **Supplementary Figure 1E**) nor within any subgroups.

**Figure 2.**
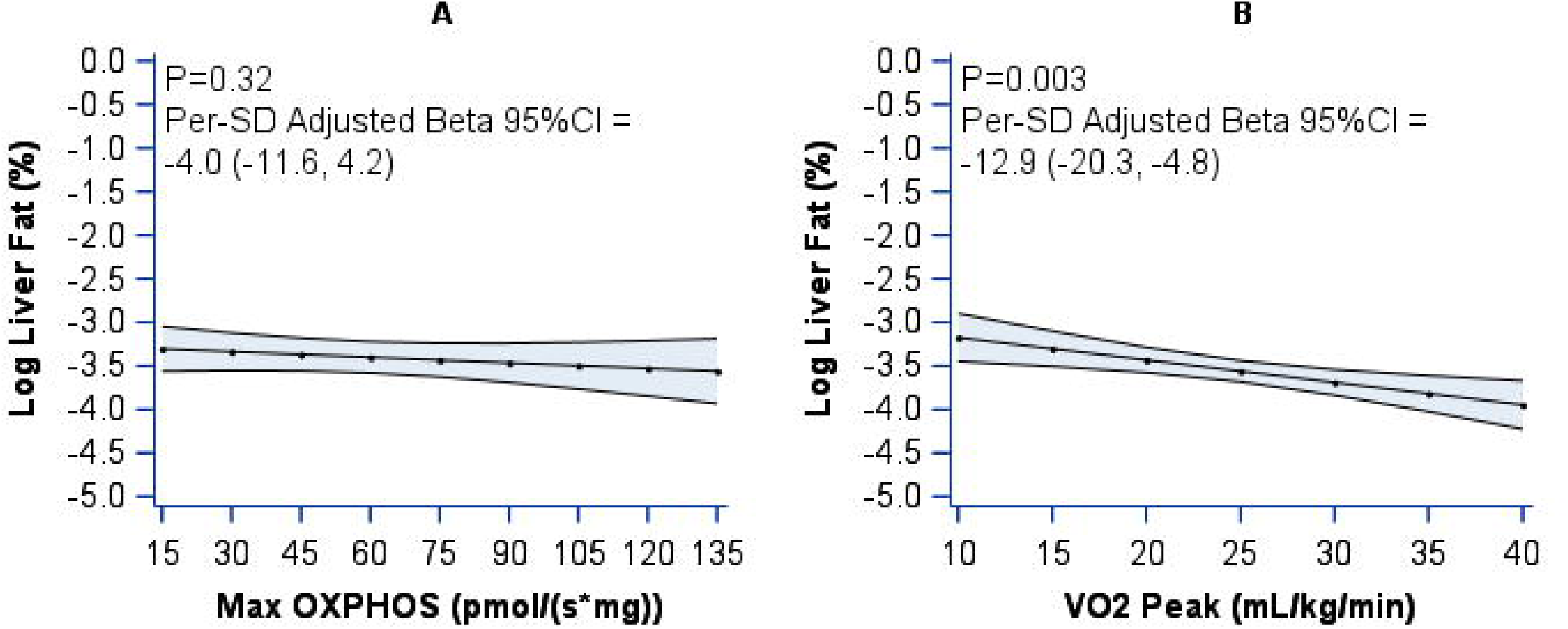
Associations of Muscle Mitochondrial Energetics and Cardiorespiratory Fitness With Liver Fat Among SOMMA Older Adults Panel (A) shows the association between carbohydrate-supported maximum oxidative phosphorylation (i.e., muscle mitochondrial energetics) and log-transformed liver fat (N=321). Panel (B) shows the association of cardiorespiratory fitness measured as VO_2_ peak with liver fat (N=361). Models were adjusted for age, gender, daily minutes of moderate to vigorous physical activity, and prediabetes/diabetes status. Abbreviations: Max OXPHOS—maximum oxidative phosphorylation; VO_2_ peak—peak oxygen uptake

### Cardiorespiratory Fitness was Associated Inversely with Liver Fat

Cardiorespiratory fitness (i.e., VO_2_ peak) is the whole-body manifestation of muscle mass and muscle mitochondrial energetics^8^ and a biomarker of health status that declines in aged populations and in individuals with NAFLD—albeit according to studies with indirect scores estimating liver fat.^24,25^ Thus, we hypothesized that in older adults, a higher VO_2_ peak would be associated with lower liver fat when using gold-standard MRI proton density fat fraction. Indeed, each SD increment in VO_2_ peak (mL/kg/min) was associated with a ∼13% lower liver fat in the fully adjusted model (**Figure 2B**, **Supplementary Figure 1F**; fully adjusted β: −12.9; 95%CI: −20.3, −4.8; *p*=.003; N=361) and did not vary across strata of gender or waist circumference.

### Skeletal Muscle-Related Characteristics are Associated with the Odds of NAFLD

The results using dichotomous NAFLD as the outcome were similar in directionality to the primary results. Specifying NAFLD dichotomously additionally indicated a trend towards a negative association between D_3_Cr muscle mass/wgt and the odds of NAFLD among participants with a normal waist circumference; however, the result did not reach statistical significance (OR: 0.65, 95%CI: 0.39, 1.1, *p*=0.11; **Supplementary Table 2**).

Contrary to the primary results, supplemental results indicated a potential interaction (*p*=.09) between ATP_max_ and gender for the odds of NAFLD. However, neither of the stratified gender-specific estimates were statistically significant. This may indicate a spurious interaction owing to a sparse data distribution at the high end of ATP_max_, which almost always corresponded with a low liver fat; **Supplementary Figure 3E**). Thus, the interaction should be interpreted with caution and confirmed in future studies with adequate power.

## Discussion

Loss of skeletal muscle mass alongside myosteatosis and decreased cardiorespiratory fitness^24,26^ are common phenotypes in aging^5^ that may also be present in NAFLD.^13^ However, associations between muscle mass and liver fat have not been replicated in all studies,^5,6^ remain understudied in older cohorts, and heretofore the associations between muscle mitochondrial energetics and liver fat remained largely unknown. Therefore, we asked whether deeply-phenotyped characteristics of skeletal muscle and cardiorespiratory fitness were associated with liver fat in older adults using state-of-the-art measurements. We did not find evidence that D_3_Cr muscle mass/wgt *per se* was associated with liver fat in our overall sample; however, the association was positive among older adults with a high waist circumference—a surrogate for visceral adipose volume.^27^ As an adjunct to these findings, our supplementary results suggest that a higher D_3_Cr muscle mass/wgt may be associated with a lower odds of the clinical NAFLD phenotype among older adults *without abdominal obesity* (i.e., with a normal waist circumference), although the lack of statistical significance necessitates confirmation of the trend in future studies. Using robust imaging techniques, we found that muscle fat infiltration was associated with liver fat in an aged population in whom maintenance of muscle health is a major concern.^28^ Importantly, our investigation illuminates a potential link between higher carbohydrate-supported *ex vivo* muscle mitochondrial energetics and lower liver fat, albeit no association between *in vivo* mitochondrial energetics and liver fat was observed. Finally, our study uncovered a robust inverse association between VO_2_ peak and liver fat in this aged study sample.

Our results that capitalized on precise measurements of both muscle mass and liver fat highlight the need to consider abdominal adiposity as a modifying factor that may alter the association between the two tissues in older adults.^5,14^ We found that muscle mass and liver fat were directly associated in older adults with a high waist circumference and who may have greater visceral adipose depots,^29^ but trended towards being inversely associated among those *without* abdominal obesity (as hypothesized). This aligns with prior literature demonstrating that visceral adiposity confers metabolic risk even at higher relative quantities of lean mass.^30^ This general principle is likely magnified among older adults in whom adiposity as a proportion of total body mass, and in particular central adiposity, are exacerbated with aging and relate to functional decline.^31^ This may be because a higher waist circumference and therefore visceral fat could theoretically drive muscle fat infiltration and loss of muscle integrity. Our results lead us to speculate that dysfunctional muscle tissue could promote liver fat accumulation via muscle insulin resistance, a frequent comorbidity of central adiposity,^32^ even in older adults with high relative muscle mass. In the context of this dysfunctional metabolic milieu, increased availability of free fatty acids from adipose tissue or other substrates such as glucose may drive *de novo* hepatic lipogenesis. In contrast, a higher relative muscle mass may be inversely associated with liver fat among older adults with a normal waist circumference *independent of reported physical activity levels* due to the maintenance of insulin sensitivity; whether molecular muscle characteristics such as mitochondrial energetics explain this trend^33^ should be elucidated in prospective studies.

One reason for the lack of consensus in the literature regarding the association between muscle mass and liver fat may be due to the use of indirect muscle and liver fat^5,34,35^ measurements or those that are complicated by changes in body water commonly present in chronic diseases and aging (e.g., bioelectrical impedance analysis^35^ or dual energy x-ray absorptiometry in the case of muscle mass^5^). The measurement methods we used circumvented these challenges. Despite having a comparable distribution of muscle mass^36^ and liver fat^37,38^ to contemporary population-based cohorts of older adults, our results may also be discordant with some prior research due to differences in participant demographics. Much of the available data describing the muscle-liver axis has been collected in younger (mean age ∼45-60 years) study samples^5,35^ comprised predominantly of participants with an Asian lineage,^5,35^ who may have noteworthy differences in regional body composition as compared to non-Hispanic White individuals (which represent the majority of SOMMA participants).^39^ Finally, given that SOMMA was not expressly designed to recruit participants across the spectrum of NAFLD severity, study participants had relatively low levels of liver fat, although this is not necessarily an indicator of NAFLD progression.^40^ We emphasize that our results should be interpreted within the context of these cross-study differences.

Our results and those of other impactful research studies strongly point to the need to look beyond muscle mass and more deeply into how specific muscle characteristics relate to liver fat. One such characteristic is muscle fat infiltration. In line with our results in older adults with a median age of 75 (range 70-93) years, muscle fat infiltration was a superior correlate of liver fat than sarcopenia (low hand grip strength and height-adjusted appendicular lean mass) before adjustment for BMI among 5,326 adults in the population-based UK Biobank study who were over a decade younger (63 ± 7.5 years), on average.^6^ Another recent report similarly found that myosteatosis (a more direct measure of intramuscular fat) is a stronger determinant of early-stage non-alcoholic steatohepatitis (i.e., a more advanced stage of NAFLD characterized by inflammation, ballooned hepatocytes, and/or fibrosis)^1^ as compared to sarcopenia among middle-aged participants.^9^ We speculate that lower muscle fat infiltration, particularly in untrained individuals, may reflect greater skeletal muscle oxidative capacity through enhanced mitochondrial function or increased glucose disposal,^5^ thereby shunting less glucose to the liver and potentially improving liver insulin sensitivity.^41^

Indeed, our study revealed for the first time, to our knowledge, that muscle oxidative capacity—a key muscle characteristic contributing to cardiometabolic risk that is diminished in aging and obesity^20^—may be an important mechanistic component of the muscle-liver fat axis in older adults. Of note, *liver* mitochondrial energetics are impaired in animal models and in humans with NAFLD,^33,42,43^ but the association of *muscle* mitochondrial energetics with liver fat has never been described in humans using standardized *ex vivo* assays. Our results suggest that higher carbohydrate-supported skeletal muscle mitochondrial respiration (i.e., Max OXPHOS) as measured *ex vivo* in permeabilized fiber bundles is associated with lower liver fat. The attenuation of this result through adjustment for potential confounding variables including prediabetes/diabetes status may point to the diabetes-related hallmark of insulin resistance as a key pathophysiologic factor linking lower mitochondrial respiration in both skeletal muscle and liver.^33,44^ Lower mitochondrial energetics and skeletal muscle insulin resistance could drive postprandial glucose flux to the liver where increased substrate-driven *de novo* lipogenesis may contribute to hepatic steatosis.^41^ In contrast to carbohydrate-supported OXPHOS, fatty acid-supported muscle mitochondrial OXPHOS was *not* correlated with liver fat. Although speculative, these results support a “reroute hypothesis,” which posits that more optimal carbohydrate-supported OXPHOS in peripheral tissues such as muscle prevents energy storage in hepatic tissues, preventing downstream liver insulin resistance and hepatic steatosis.^22^ This should be explored further in studies designed for this purpose and in individuals with diabetes specifically.

In contrast to our *ex vivo* measure of mitochondrial energetics (Max OXPHOS), we did not find a statistically significant association between ATP_max_–our secondary measure of mitochondrial energetics taken *in vivo*—and liver fat in our sample of older adults. This negative result recapitulates the finding that *in vivo* mitochondrial function measured by ^31^P-MRS of the quadriceps muscle was not different in 17 participants with NAFLD compared with 18 age- and BMI-matched controls.^11^ This may be due to interindividual variability in blood flow or local substrate levels; or in the delivery, transport, and metabolism of substrates needed to fuel ATP_max_ that are standardized in the *ex vivo* respirometry assay.^45^ Thus, the standardization procedures conducted in the *ex vivo* assay may be necessary to observe potential linkages between muscle mitochondrial energetics and liver fat.

We and others have shown that oxidative capacity within the muscle (i.e., muscle mitochondrial energetics) is related to whole-body oxygen consumption (i.e., cardiorespiratory fitness measured as VO_2_ peak), muscle mass, and whole-body metabolic health among older adults.^8,46^ Building on these findings, our exploratory analyses uncovered an inverse association between VO_2_ peak and liver fat among our older adult study sample, which was robust to adjustment for confounding variables including prediabetes/diabetes status. This result echoes prior literature conducted in younger study samples, including an interventional study in which results withstood adjustment for fat mass loss, despite VO_2_ peak estimation using cycle ergometry (as opposed to CPET—the gold-standard measurement used in our study).^47^ Future studies should investigate whether VO_2_ peak mediates the association between muscle mitochondrial energetics with liver fat, as an enhanced oxidative capacity of muscle mitochondria could theoretically prevent ectopic fat accumulation in the liver while raising VO_2_ peak.^46,48^ Furthermore, VO_2_ peak, mitochondrial energetics, and functional measures such as walk speed are closely related,^8^ so elucidating the causal pathway by which these variables relate to liver fat will be key to developing interventions that address both liver fat and functional outcomes in older adults. Taken together, these findings support the premise that specific muscle characteristics and associated phenotypes may be key for understanding muscle-liver crosstalk, particularly in the aging population in whom central adiposity may render muscle mass less effective at protecting against liver fat accumulation. Central adiposity appears to be a relevant modifier of the association between skeletal muscle mass and liver fat, but not for associations of muscle-related characteristics and liver fat. These hypothesis-generating results require confirmation in prospective studies.

This study includes several limitations and strengths. We were unable to test the directionality of the muscle-liver association in our cross-sectional study, which is important to consider given that the liver communicates with muscle tissue via circulating hepatokines,^49^ delivers excess lipid to muscle via lipoproteins, and can contribute to muscle inflammation that in turn drives sarcopenia.^5,50^ Indeed, people with advanced stages of NAFLD, such as fibrosis and cirrhosis, often have reduced muscle mass and impaired muscle function^51^ and these characteristics are associated with poor prognosis (e.g., hospitalizations, ascites, and encephalopathy).^52^ Directionality can be evaluated in future studies with collection of longitudinal data. Our study was underpowered to test for interaction between Max OXPHOS and diabetes status for the outcome of liver fat, which may be a critical element to include in follow-up research to elucidate mechanisms of muscle-liver crosstalk. Although unlikely, some participants who self-reported as having diabetes may have had type 1 diabetes, in whom mechanisms linking muscle and liver phenotypes may be unique. Our analysis is subject to residual confounding as is the case with virtually all observational epidemiologic studies. The primary strength of this study is the rigor and breadth of our measurement methods for determining muscle characteristics and liver fat content in an aged cohort, which gives rise to hypotheses about mechanisms liking skeletal muscle mitochondrial energetics and liver fat.

In conclusion, our results reveal muscle characteristics that are associated with liver fat in older adults, suggesting that muscle and liver health are linked potentially via specific mechanisms and biomarkers. Our finding that both muscle mitochondrial bioenergetics and cardiorespiratory fitness are associated with liver fat accumulation suggests that clinical assessments of fitness, coupled with standard biochemical measures, could provide a scalable approach to evaluate risk factors for NAFLD in the aging population. Future studies with prospective designs should confirm and expand upon these results using both lifestyle and pharmacological interventions. In particular, exercise interventions may serve as a model for studying the complex links amongst muscle characteristics and liver fat in older adults as exercise increases cardiorespiratory fitness, muscle mass,^53^ and muscle mitochondrial respiration^13^ while reducing liver fat. Such study designs will enrich our knowledge of the directionality and mechanistic underpinnings of the muscle-liver axis and ultimately inform optimization of NAFLD treatment and prevention in the aging population.^54^

## Supporting information

Graphical abstract

## List of Abbreviations

NAFLD: non-alcoholic fatty liver disease
SOMMA: Study of Muscle, Mobility and Aging
MRS: magnetic resonance spectroscopy
BMI: body mass index
CPET: Cardiopulmonary Exercise Testing
Max OXPHOS: maximal oxidative phosphorylation
ATP_max_: maximum adenosine triphosphate production

## Acknowledgements

We acknowledge all SOMMA staff and investigators and thank all SOMMA participants who enabled this research.

## Author contributions

Paul M. Coen, Karen D. Corbin, and Daria Igudesman contributed substantially to the conception and design of the manuscript. Daria Igudesman, Justine Mucinski, Paul M. Coen, and Karen D. Corbin wrote the original draft. Daria Igudesman conducted the formal analysis and curated the data with input from Stephanie Harrison, Peggy M. Cawthon, and Jennifer Linge. Paul M. Coen, Peggy M. Cawthon, Bret H. Goodpaster, Steven R. Cummings, Russell T. Hepple, Stephanie Harrison, Jennifer Linge, and Stephen B. Kritchevsky were involved in SOMMA study investigation. Steven R. Cummings, Russell T. Hepple, Stephen B. Kritchevsky, and Anne B. Newman acquired funding. All study authors, including Michael J. Jurczak and David Marcinek, participated in extensive review and editing of the manuscript.

## Data availability

The data that support the findings of this study are available from the Study of Muscle, Mobility and Aging (SOMMA). As of August 10^th^, 2023, data can be obtained via https://sommaonline.ucsf.edu

## Ethics Approval

All research was conducted in accordance with the Declaration of Helsinki. All participants provided written informed consent, and the study was approved by the WIRB-Copernicus Group Institutional Review Board (20180764) for all participating sites.

## Participant Informed Consent

All patients provided written informed consent for the anonymous use of their data for research purpose.

