## Supplementary material for "Associations of Skeletal Muscle Mass, Muscle Fat Infiltration, Mitochondrial Energetics, and Cardiorespiratory Fitness with Liver Fat Among Older Adults": Graphical abstract

### Slide 1
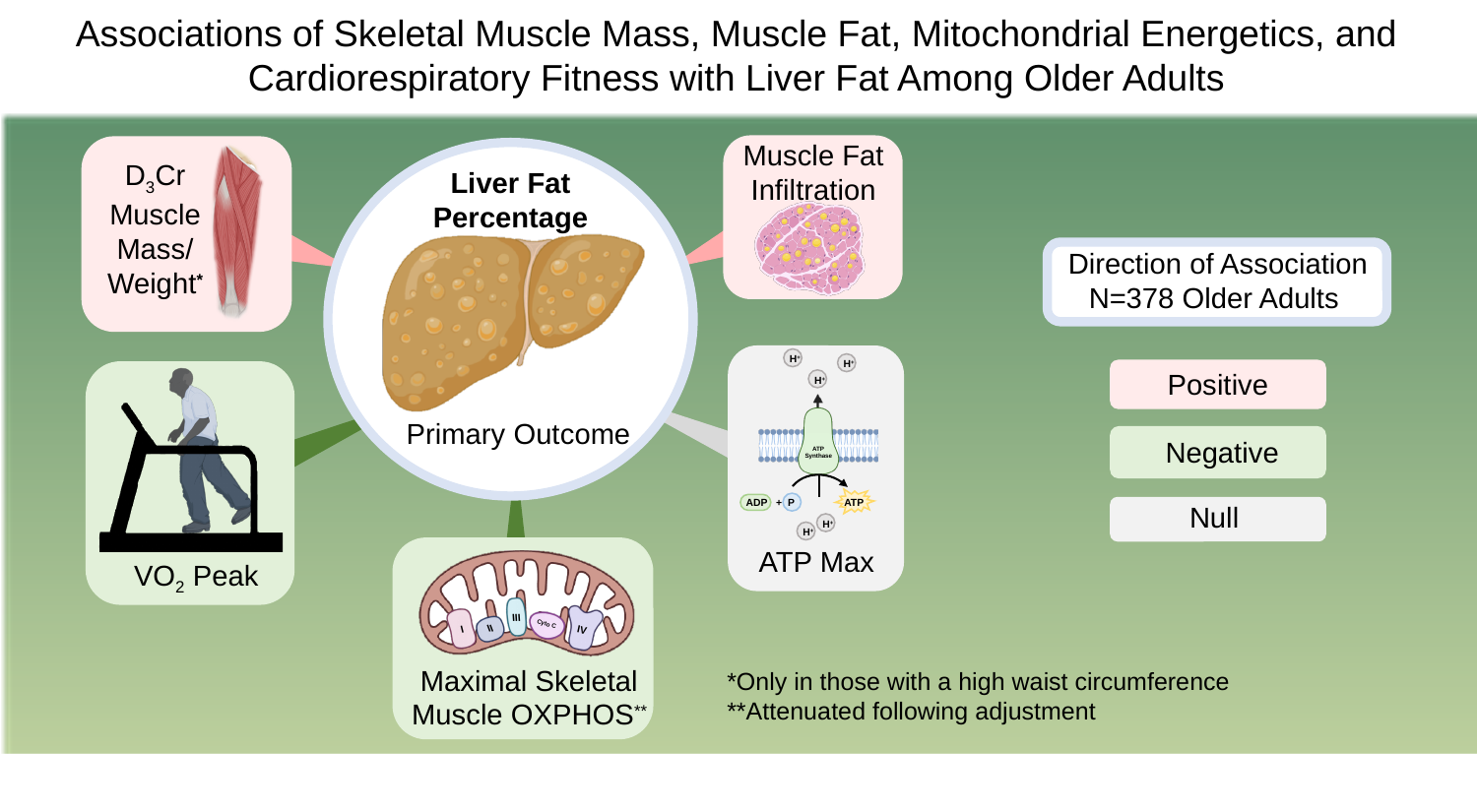

Associations of Skeletal Muscle Mass, Muscle Fat, Mitochondrial Energetics, and Cardiorespiratory Fitness with Liver Fat Among Older Adults
Muscle Fat Infiltration
D_3_Cr Muscle Mass/
Weight^*^
Primary Outcome
Liver Fat Percentage
Direction of Association N=378 Older Adults
Positive
Negative
Null
H^+^
H^+^
H^+^
ATP
Synthase
ATP
ADP
P
+
H^+^
H^+^
ATP Max
VO_2_ Peak
III
Cyto C
II
I
IV
Maximal Skeletal Muscle OXPHOS^**^
*Only in those with a high waist circumference
**Attenuated following adjustment
