## Supplemental data for "Associations of Skeletal Muscle Mass, Muscle Fat Infiltration, Mitochondrial Energetics, and Cardiorespiratory Fitness with Liver Fat Among Older Adults"

**Supporting Information**

**Supplementary Table 1. Comparison of Baseline Characteristics of Participants Included and Excluded from Analysis**

**Supplementary Table 2. Associations of D3Cr Muscle Mass/Wgt, Muscle Characteristics, and Cardiorespiratory Fitness with the Odds of Non-Alcoholic Fatty Liver Disease**

**Supplementary Figure 1. Participant Flow Diagram**

**Supplementary Figure 2. Sequentially Adjusted Models for Primary Analysis of D_3_Cr Muscle Mass/Wgt and Liver Fat Percentage**

**Supplementary Figure 3. Crude Scatterplots of D_3_Cr Muscle Mass/Wgt, Muscle Characteristics, and Cardiorespiratory Fitness with Liver Fat Percentage**

| **Supplementary Table 1. Comparison of Baseline Characteristics of Participants Included and Excluded from Analysis** | | | | | |
| --- | --- | --- | --- | --- | --- |
|  | **Included** | | **Excluded** | |  |
|  | **Total N** | **Mean ± SD, Median (IQR), or N (%)** | **Total N** | **Mean ± SD, Median (IQR), or N (%)** | **P-value** |
| Age (years) | 378 | 75 (73, 79) | 501 | 75 (72, 79) | 0.13 |
| Women | 378 | 226 (59.8) | 501 | 294 (58.7) | 0.74 |
| Race | 378 |  | 501 |  | 0.30 |
| Asian |  | <5 |  | <5 |  |
| Black or African  American |  | 45 (11.9) |  | 71 (14.2) |  |
| Native American/  Alaskan Native |  | <5 |  | <5 |  |
| Multiracial |  | 5 (1.0) |  | <5 |  |
| Other race/unknown |  | <5 |  | <5 |  |
| White or Caucasian |  | 327 (86.5) |  | 418 (83.4) |  |
| Hispanic Ethnicity | 378 | <5 | 501 | 6 (1.2) | 0.74 |
| BMI (kg/m²) | 378 | 27.8 ± 4.8 | 501 | 27.5 ± 4.4 | 0.32 |
| Waist circumference | 372 |  | 482 |  | **0.04** |
| Normal |  | 205 (55.1) |  | 232 (48.1) |  |
| High |  | 167 (44.9) |  | 250 (51.9) |  |
| Prediabetes or diabetes | 378 | 141 (37.3) | 500 | 236 (47.2) | **0.003** |
| Hemoglobin A1c (%) | 373 | 5.5 (5.3, 5.8) | 492 | 5.6 (5.4, 5.9) | **0.0001** |
| Moderate to vigorous Physical activity (daily minutes) | 378 | 188 ± 87 | 444 | 184 ± 85 | 0.51 |
| Liver fat percentage (%) | 378 | 2.8 (2.0, 5.4) | 52 | 3.1 (2.2, 5.1) | 0.47 |
| D_3_Cr muscle mass/wgt (%) | 362 | 28.7 ± 6.8 | 468 | 29.3 ± 6.5 | 0.16 |
| Muscle fat infiltration (%) | 378 | 7.3 (6.0, 9.1) | 455 | 6.4 (5.1, 7.8) | **<0.0001** |
| Maximal carbohydrate-supported oxidative phosphorylation (pmol/(s*mg)) | 321 | 59.7 ± 19.2 | 424 | 59.7 ± 17.9 | 0.99 |
| ATP Production (mM/sec) | 345 | 0.52 ± 0.14 | 467 | 0.56 ± 0.15 | **0.0004** |
| VO_2_ peak (mg/kg/min) | 361 | 20.9 ± 5.4 | 459 | 19.7 ± 4.3 | **0.0003** |
| Continuous variables presented as mean ± SD or median (IQR). Categorical variables presented as N (%). Prediabetes or diabetes ascertained based on HbA1c (≥5.7%) or self-report of diabetes. A normal waist circumference was considered to be <88 cm for women and <102 cm for men. | | | | | |

| **Supplementary Table 2. Associations of D_3_Cr Muscle Mass/Wgt, Muscle Characteristics, and Cardiorespiratory Fitness with the Odds of Non-Alcoholic Fatty Liver Disease** | | | | |
| --- | --- | --- | --- | --- |
| **Exposure Variables** | **N** | **SD** | **NAFLD**  **OR (95%CI)** | **P-value** |
| Primary Exposure Variable |  |  |  |  |
| D_3_Cr Muscle Mass/Wgt |  |  |  |  |
| Model 1 | 362 | 6.8 | 0.92 (0.72, 1.2) | 0.48 |
| Model 2 | 362 | 6.8 | 0.93 (0.71, 1.2) | 0.58 |
| Model 3 | 362 | 6.8 | 1.02 (0.77, 1.3) | 0.91 |
| Waist Circumference Interaction† | 357 | -- | -- | **0.003** |
| Normal | 197 | 6.7 | 0.65 (0.39, 1.1) | 0.11 |
| High | 160 | 5.6 | 2.1 (1.4, 3.2) | **0.0004** |
| Gender Interaction | 362 | -- | -- | 0.70 |
| Exploratory Exposure Variables |  |  |  |  |
| Muscle Fat Infiltration |  |  |  |  |
| Model 1 | 378 | 2.3 | 1.6 (1.3, 2.0) | **<0.0001** |
| Model 2 | 378 | 2.3 | 1.7 (1.3, 2.2) | **<0.0001** |
| Model 3 | 378 | 2.3 | 1.6 (1.2, 2.0) | **0.001** |
| Waist Circumference Interaction† | 372 | -- | -- | 0.47 |
| Gender Interaction | 378 | -- | -- | 0.90 |
| Carbohydrate-Supported Maximum  Oxidative Phosphorylation  (pmol/(s*mg)) |  |  |  |  |
| Model 1 | 321 | 19.2 | 0.88 (0.68, 1.1) | 0.34 |
| Model 2 | 321 | 19.2 | 0.96 (0.72, 1.3) | 0.80 |
| Model 3 | 321 | 19.2 | 1.04 (0.77, 1.4) | 0.81 |
| Waist Circumference Interaction† | 321 | -- | -- | 0.39 |
| Gender Interaction | 321 | -- | -- | 0.21 |
| ATP Production (mM/sec) |  |  |  |  |
| Model 1 | 345 | 0.14 | 0.88 (0.68, 1.1) | 0.32 |
| Model 2 | 345 | 0.14 | 0.88 (0.68, 1.1) | 0.80 |
| Model 3 | 345 | 0.14 | 0.98 (0.74, 1.3) | 0.87 |
| Waist Circumference Interaction† | 339 | -- | -- | 0.41 |
| Gender Interaction | 345 | -- | -- | **0.09** |
| Women | 209 | 0.14 | 0.43 (0.10, 1.8) | 0.25 |
| Men | 136 | 0.15 | 2.1 (0.48, 9.5) | 0.32 |
| VO_2_ peak (mL/kg/min) |  |  |  |  |
| Model 1 | 361 | 5.4 | 0.60 (0.45, 0.80) | **0.0005** |
| Model 2 | 361 | 5.4 | 0.53 (0.38, 0.74) | **0.0002** |
| Model 3 | 361 | 5.4 | 0.57 (0.40, 0.83) | **0.003** |
| Waist Circumference Interaction† | 355 | -- | -- | 0.81 |
| Gender Interaction | 361 | -- | -- | 0.45 |
| Model 1. Crude  Model 2. Adjusted for age, gender (model with maximum oxidative phosphorylation further adjusted for respirometry technician)  Model 3. Further adjusted for daily minutes of MVPA and prediabetes/diabetes status  The back-transformed beta estimate represents the relative percentage difference in liver fat associated with a 1-SD increment in each independent variable. Interaction models are fully adjusted. Stratified estimates are shown when the p-value for interaction was <0.1.  †Normal waist circumference was defined as <88 cm for women and <102 cm for men.  Abbreviations: NAFLD—non-alcoholic fatty liver disease | | | | |

**Supplementary Figure 1.** Participant Flow Diagram

378 Older Adults Included in Analysis of Muscle Fat Infiltration

321 with Max OXPHOS

57 excluded due to missing Max OXPHOS

345 with ATP Max

33 excluded due to missing ATP Max

16 excluded due to missing muscle mass data

362 with D_3_Cr Muscle Mass/Wgt

17 excluded due to missing VO_2_ peak data

361 with VO_2_ peak

37 excluded due to missing physical activity data

415 Older Adults Enrolled at Pittsburg Site with Liver Fat Data

879 Older Adults Enrolled in SOMMA

- 449 excluded due to missing liver fat data
- 15 excluded from Wake Forest site

Abbreviations: SOMMA—Study of Muscle, Mobility, and Ageing; Max OXPHOS—maximum oxidative phosphorylation

**Supplementary Figure 2. Sequentially Adjusted Models for Primary Analysis of D_3_Cr Muscle Mass/Wgt and Liver Fat Percentage**

Gender

Age

Model 2

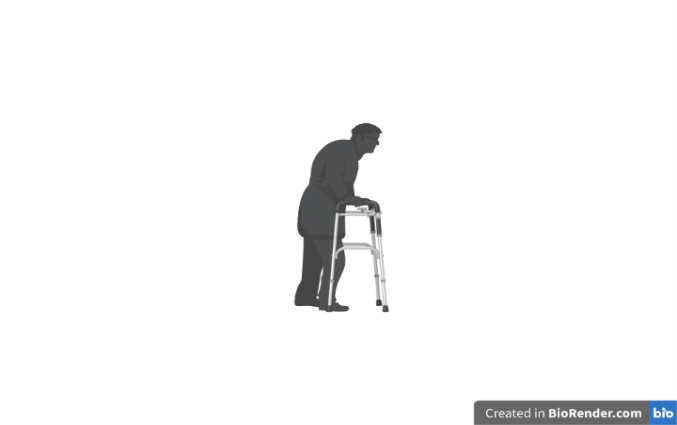

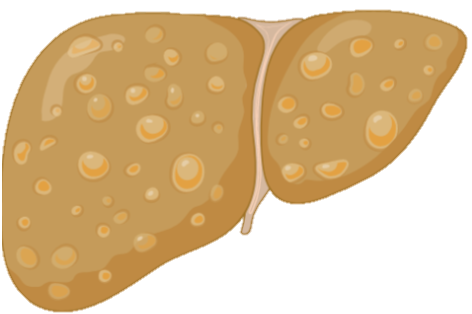

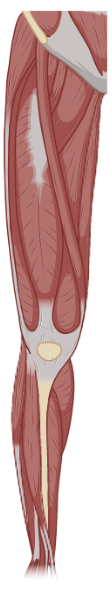

**Primary Outcome**

Liver Fat Percentage

**Primary Exposure**

D_3_Cr Muscle Mass Percentage

Model 1 (Crude)

Model 3

+ MVPA

Effect Modifiers

Model 3b

Gender

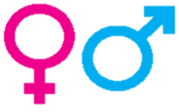

Model 3a

Waist Circumference

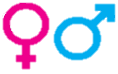

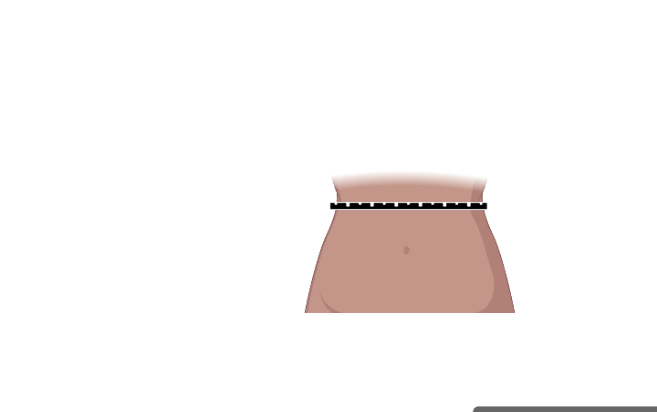

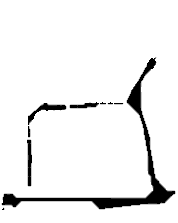

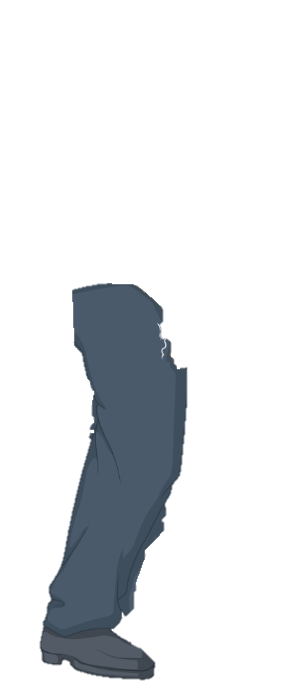

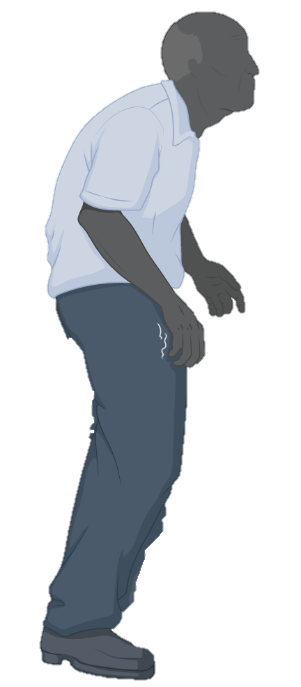

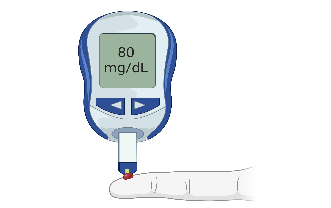

+Prediabetes/

Diabetes Status

Effect modifiers were included in interaction terms with the exposure of D_3_Cr muscle mass percentage in fully adjusted models. Models 2-3b included technician when maximum oxidative phosphorylation (respirometry) was the independent variable. Abbreviations: MVPA—moderate to vigorous physical activity; Wgt—Weight

**Supplementary Figure 3.** Crude Scatterplots of D_3_Cr Muscle Mass/Wgt, Muscle Characteristics, and Cardiorespiratory Fitness with Liver Fat Percentage

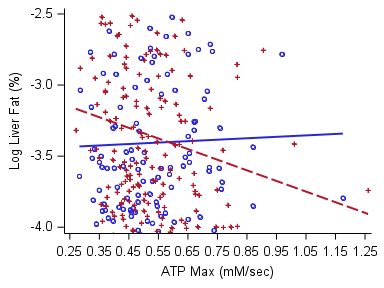

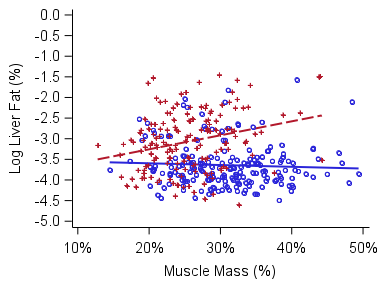

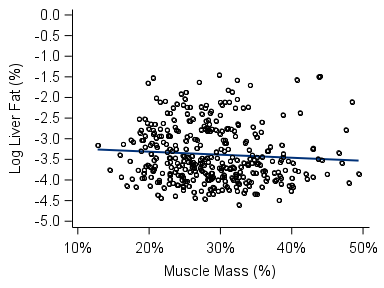

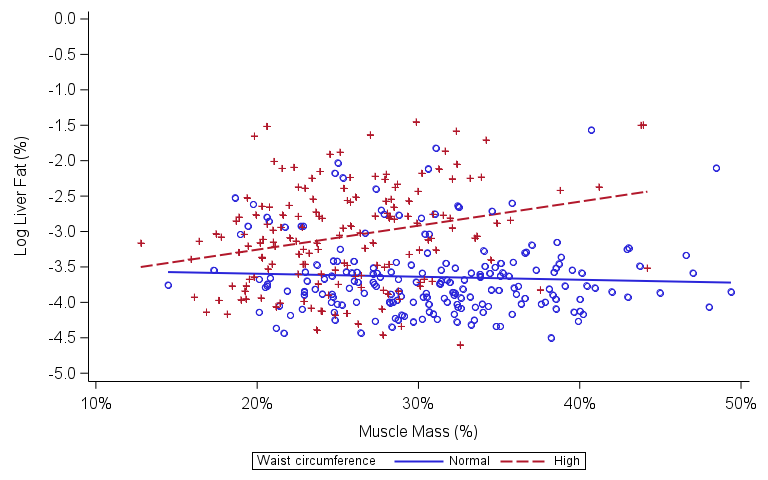

A

B

C

D

E

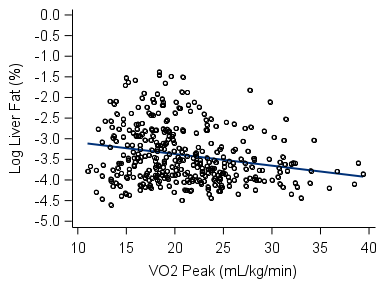

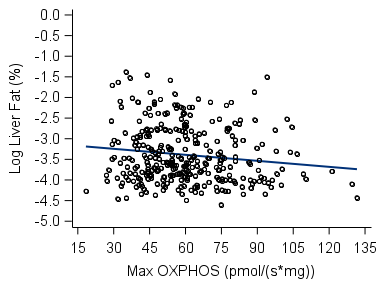

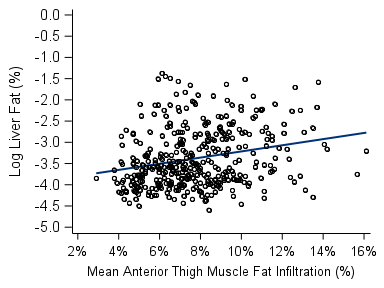

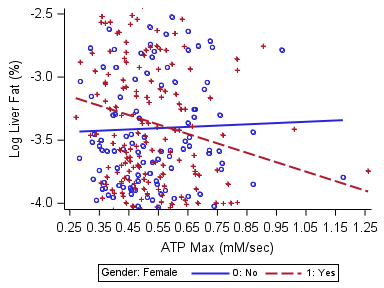

F

Raw, unadjusted scatterplots of muscle mass/wgt with liver fat percentage overall (Panel A, *N* = 362) and stratified by waist circumference (Panel B, *N* = 357). Panels C-E show raw scatterplots of exploratory independent variables (muscle fat infiltration, *N* = 378; maximal carbohydrate-supported oxidative phosphorylation, *N* = 321; and VO_2_ peak, *N* = 361; respectively) with the outcome of liver fat percentage. A normal waist circumference was considered to be <88 cm for women and <102 cm for men. Abbreviations: Max OXPHOS—maximum oxidative phosphorylation**;** VO_2_ peak—peak oxygen uptake; Wgt—weight
